# Reconciling model predictions with low reported cases of COVID-19 in Sub-Saharan Africa: Insights from Madagascar

**DOI:** 10.1101/2020.07.15.20149195

**Authors:** Michelle V. Evans, Andres Garchitorena, Rado J.L. Rakotonanahary, John M. Drake, Benjamin Andriamihaja, Elinambinina Rajaonarifara, Calistus N. Ngonghala, Benjamin Roche, Matthew H. Bonds, Julio Rakotonirina

## Abstract

The COVID-19 pandemic has wreaked havoc globally, and there has been a particular concern for sub-Saharan Africa (SSA), where models suggest that the majority of the population will become infected. Conventional wisdom suggests that the continent will bear a higher burden of COVID-19 for the same reasons it suffers high burdens of other infectious diseases: ecology, socio-economic conditions, lack of water and sanitation infrastructure, and weak health systems. However, so far SSA has reported lower incidence and fatalities compared to the predictions of standard models and the experience of other regions of the world. There are three leading explanations, each with very different implications for the final epidemic burden: (1) low case detection, (2) differences in COVID-19 epidemiology (e.g. low *R*_*0*_), and (3) policy interventions. The low number of cases to date have led some SSA governments to relax these policy interventions. Will this result in a resurgence of cases? To understand how to interpret the lower-than-expected COVID-19 case data in Madagascar, we use a simple age-structured model to explore each of these explanations and predict the epidemic impact associated with them. We show that the current incidence of COVID-19 cases can be explained by any combination of the late introduction of first imported cases, early implementation of non-pharmaceutical interventions (NPIs), and low case detection rates. This analysis reinforces that Madagascar, along with other countries in SSA, remains at risk of an impending health crisis. If NPIs remain enforced, up to 50,000 lives may be saved. Even with NPIs, without vaccines and new therapies, COVID-19 could infect up to 30% of the population, making it the largest public health threat in Madagascar until early 2021, hence the importance of conducting clinical trials and continually improving access to healthcare.

**Résumé:** La pandémie de COVID-19 a eu des conséquences néfastes partout dans le monde, et il y a une préoccupation particulière pour l’Afrique subsaharienne (ASS), où des modèles suggèrent que la majorité de la population sera infectée. Il est craint que le continent supportera un fardeau plus élevée de COVID-19 pour les mêmes raisons qu’il souffre d’avantage d’autres maladies infectieuses: écologie, conditions socio-économiques, manque d’infrastructures d’eau et d’assainissement, et faiblesse des systèmes de santé. Cependant, jusqu’à présent, l’ASS a rapporté une incidence et une mortalité bien inférieure à celle des prévisions des modèles pour cette région, ainsi qu’au nombre observé dans d’autres régions du monde. Il y a trois explications principales pour ce phénomène, chacune ayant des implications très différentes pour le fardeau épidémique final: (1) détection faible des cas, (2) différences dans l’épidémiologie COVID-19 (par exemple faible R0), et (3) interventions et politiques mises en place. Le faible nombre de cas à ce jour a conduit certains gouvernements d’ASS à assouplir ces interventions. Cela entraînera-t-il une résurgence de cas? Pour comprendre comment interpréter le fait que les cas COVID-19 rapportés sont plus faibles que prévu à Madagascar, nous utilisons un modèle de transmission structuré par groupe d’âge pour explorer chacune de ces explications et prédire l’impact épidémique qui leur est associé. Nous montrons que l’incidence actuelle des cas de COVID-19 peut s’expliquer par l’effet cumulé de l’introduction tardive des premiers cas importés, la mise en œuvre rapide d’interventions non pharmaceutiques (INP) et de faibles taux de détection des cas. Cette analyse renforce le fait que Madagascar, ainsi que d’autres pays d’Afrique subsaharienne, reste à risque d’une crise sanitaire imminente. Si les INP restent appliqués, jusqu’à 50 000 vies pourraient être sauvées. Même avec des INP, tant qu’il n’y aura pas des vaccins ni des nouvelles thérapies efficaces, COVID-19 pourrait infecter jusqu’à 30% de la population, ce qui constituerait la plus grande menace pour la santé publique à Madagascar jusqu’au début de 2021, d’où l’importance de la réalisation des essais cliniques et de l’amélioration continuelle de l’accès aux soins.

**Summary Box:**

- The lower-than-expected number of reported cases of COVID-19 in Madagascar can be explained by a combination of the relatively late introduction of the disease, low detection rates, and low transmission rates due to the early and effective implementation of non-pharmaceutical interventions that reduced contact rates.
- COVID-19 is predicted to be the largest public health problem in Madagascar in 2020, but non-pharmaceutical interventions at an effectiveness similar to those seen in the first few months could save up to 50,000 lives.
- Health systems in SSA remain at risk of an impending health crisis due to COVID-19, stressing the importance of ongoing clinical trials and improving health care access.

## Introduction

The COVID-19 pandemic has killed hundreds of thousands of people, collapsing health systems and economies around the world. Most models predict that without intervention, the majority of the global population will become infected and tens of millions will die as a result of the pandemic [1]. There have been particular concerns for sub-Saharan Africa (SSA) [2–4], as the major factors that drive high burdens of other infectious diseases, such as the environmental and socio-economic conditions, lack of water and sanitation infrastructure, and weak health systems, are equally relevant to the threat of COVID-19. However, so far, the perceived burden of COVID-19 in SSA is low compared to expectations both from epidemiological models and from epidemic patterns in other regions of the world [5,6]. Though SSA comprises 11% of the global population, it has only 3.6% of the total global COVID-19 incidence, much of which is due to case reports from South Africa [7]. As of July 2020, most SSA countries are reporting fewer than 100 new cases daily [8]. There are three leading potential explanations for the lower observed burden of COVID-19 in SSA: 1) low case detection, 2) region-specific epidemiology (e.g., different *R*_0_), and 3) early implementation of effective policy interventions. The important difference among these alternative explanations is that explanations based on low case detection and effective interventions imply that there will be a major resurgence if interventions are relaxed, while explanations based on region-specific epidemiology allow for a safe reopening.

The lower-than-expected number of reported cases may be due to low detection and reporting rates. RT-PCR laboratory capacity in SSA is limited [9] and many countries have among the lowest testing rates in the world [8]. Moreover, health care access for fever and respiratory infections is low [10], which means that many symptomatic cases will not be detected, and the stigma associated with COVID-19 could further reduce health-seeking behaviors [11].

The epidemiology-based explanations for low COVID-19 cases are based on considerations of well-established factors: warmer climates, younger age distributions, and lower contact rates due to lower population density and transportation infrastructure in rural areas [12,13]. In addition, there is considerable interest in the potential immune-mediated consequences from living in a system with greater exposure to other infectious diseases and related prophylaxis and therapeutics [14]. For example, there are major trials underway on the effects of trained immunity due to the BCG vaccine, which may increase innate immunity against a range of respiratory infections [15]. However, many of these hypotheses have recently come into doubt. The pandemic phase of COVID-19 is driven by high susceptibility, not climate [16], suggesting that warmer, humid climates will not decrease transmission at this time. Further, past outbreaks of influenza, including the 1968 pandemic and 2009 H1N1 outbreak, spread throughout the African continent and were not limited by sparse transportation networks [17]. Explanations based in region-specific epidemiology are therefore only weakly supported.

The policy response in Africa has also been a source of considerable optimism [5,18]. African governments implemented early and strong non-pharmaceutical intervention (NPI) policies that may have effectively contained disease transmission [5,19]. The first case of COVID-19 was reported in SSA one month later than the first cases in Europe, allowing countries to prepare and implement NPIs, particularly lockdowns, social distancing, masks, and regulated domestic travel, during the early stages of the pandemic [3,19,20]. Beginning in June, several SSA countries began relaxing lockdown NPIs in response to the economic and social costs of lockdown given the low reported case numbers. As partial lockdowns have been lifted, some countries’ case rates have remained stable, while others have begun to increase, leading to the WHO to urge caution and emphasize the need for a gradual and conservative release of confinement measures in SSA [21].

It remains unknown whether SSA-specific conditions will result in different epidemic dynamics in SSA than elsewhere, and whether the current lower-than-expected case burdens can be explained solely by detection rates and policies. To explore these issues, we compare COVID-19 reported case data with predictions from a simple *SEIR* compartmental model for Madagascar that integrates age-structured social contact matrices and fatality rates, assuming an *R*_*0*_ of 2.5 [22–24] (Appendix I). We then consider what levels of detection or NPI effectiveness could explain the current state of the epidemic, and whether those levels are plausible given Madagascar’s policies, demographics, and environmental context. Finally, based on these explanations, we investigate possible transmission scenarios for the first year of the epidemic, to examine the future of COVID-19 dynamics and control in Madagascar, which could be applicable to other SSA countries.

Madagascar’s demographic, economic, and health system profile is comparable to many other SSA countries [23,25,26]. It shares most of the major infectious diseases of mainland Africa (e.g. tuberculosis, malaria, respiratory infections, diarrheal diseases), and has recently endured among the worst epidemics of plague and measles in decades [27,28]. Like other SSA countries, Madagascar reported its first imported case relatively late, on March 20, 2020, and the government implemented NPIs early in the epidemic (Table 1). Madagascar instituted a national lockdown on March 23, 2020, three days before its first case attributed to local transmission. Testing practices are also similar to those in other SSA countries, initially focusing on screening for imported cases and eventually expanding to test contacts of known cases for local transmission.

**Table 1.** Timeline of non-pharmaceutical interventions (NPIs) implemented in Madagascar.

| Date | Policy / Intervention / Program | Geographic extent (Region) |
| --- | --- | --- |
| By March 19 | Allowing the remaining people outside of the country and willing to return to Madagascar to come back | National level |
| March 20 | Beginning of the epidemic in Madagascar with 3 initial imported cases announced | Analamanga* |
| March 20 | Interruption of all international and regional flights from the outside of the country | National level |
| March 21 | Following-up and testing all passengers entering Madagascar for the last 14 days for COVID-19 | National level |
| March 23 | Lockdown; curfew; interruption of all public transportation (ground and air travel) connecting Antananarivo and Toamasina to the other regions with establishment of health barrier at all national roads; prohibition of all meeting for more than 50 individuals | Analamanga and Atsinanana** |
| March 26 | Reception of 20000 Antibody RDT kits for COVID-19 testing | NA |
| March 31 to April 02 | Mass Antibody RDT testing for all passengers entering Madagascar on March 11-19 | Analamanga and Atsinanana |
| April 03 | Adding the lockdown to Fianarantsoa after detection of COVID-19 confirmed cases | Haute-Matsiatra***, Analamanga and Atsinanana |
| April 05 | Continuing lockdown and curfew | Analamanga, Atsinanana and Haute-Matsiatra |
| April 07-09 | Temporary opening of national transportation by taxi-brousse to allow people from the other regions but blocked in other cities to return home | National level |
| April 17 | Partial lifting of lockdown, which allow people to go out, as well as taxi and bus to work from 6:00 a.m. to 1:00 p.m; curfew maintained; social distancing and mandatory wearing of mask for all person going out | Analamanga, Atsinanana and Haute-Matsiatra |
| April 20 | Launching the COVID Organics tisane based on Artemisia | NA |
| April 22 | All classes preparing official exam resume | National level |
| May 04 | All previous measures maintained, and adding the Alaotra-Mangoro region; church could receive no more than 50 persons | Analamanga, Atsinanana, Haute-Matsiatra and Alaotra-Mangoro**** |
| May 04 | Church could receive no more than 200 persons | The remaining 18 regions |
| May 17 | All previous measures maintained | Analamanga, Atsinanana, Haute-Matsiatra and Alaotra-Mangoro |
| May 31 | Progressive lifting of lockdown allowing people to work from 6:00 a.m to 3:00 p.m | Analamanga |
| May 31 | Toamasina region totally locked to the other location, all classes interrupted, and people can work from 6:00 a.m. to 1:00 p.m | Atsinanana |
| May 31 | All previous measures maintained | Alaotra-Mangoro |
| May 31 | Daily life return to the normal because COVID-19 is controlled in Fianarantsoa | Haute-Matsiatra |
| June 14 | Progressive lifting of lockdown allowing people to work from 6:00 a.m to 5:00 p.m, and public transportation until 7:00 p.m | Analamanga |
| June 14 | Progressive lifting of lockdown from 6:00 a.m to 3:00 p.m | Atsinanana and Alaotra-Mangoro |

\* (Antananarivo)
\*\* (Toamasina)
\*\*\* (Fianarantsoa)
\*\*\*\* (Moramanga, Ambatondrazaka, Anosibe an'Ala)

Our exercise shows that the current incidence of COVID-19 in Madagascar can be explained by the early and effective implementation of NPIs and low case detection rates, both of which are supported by strong anecdotal evidence. In contrast, arguments of regional-specific epidemiology are based on correlational observations that have yet to be proven. This suggests that the epidemic will grow in Madagascar, and similar countries in SSA, and that these populations remain at risk of an impending health crisis. Our model indicates that, if NPIs remain enforced at the level needed to explain current case burdens, nearly 50,000 lives could be saved. Even with NPIs, 30% of the Malagasy population would become infected by March 2021, making COVID-19 the leading killer in Madagascar over this epidemic period, hence the importance of conducting clinical trials and continually improving access to healthcare.

### Case Detection

By July 2020, the simple forecast for an unmitigated epidemic predicts a daily incidence of 34,322 cases, which is nearly 500 times the reported daily incidence (Fig. 1A). Simply accounting for detection rates between 0.1 - 1% results in predictions that closely approximate the reported daily incidence of COVID-19 cases in Madagascar (Fig. 1B). Are these low levels of case detection reasonable? For countries where per capita testing is over 100-fold higher than in Madagascar (currently 79.1/100,000 population), it is estimated that less than 10% of COVID-19 cases have been detected [29]. Though the precise case detection rates for Madagascar cannot be discerned from available data, there are a number of indicators suggesting that these are lower than the already low rates of Europe or the US.

**Figure 1.**
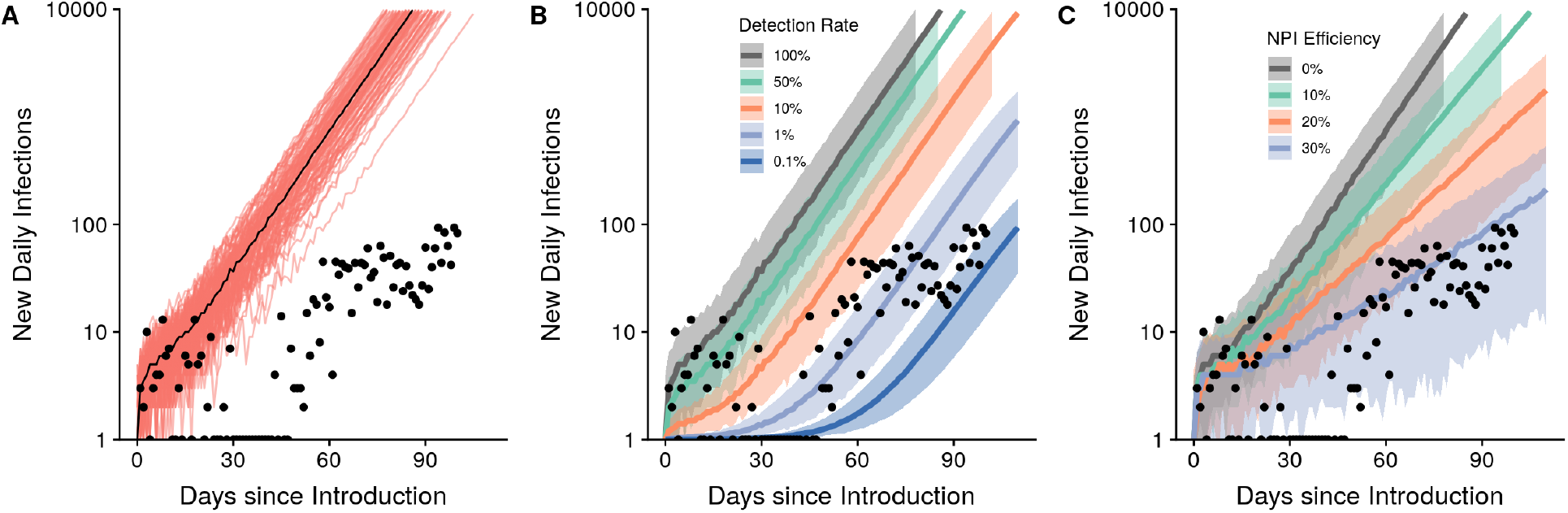
The lower-than-expected daily incidence can be explained by detection rates of 0.1-1% or NPI efficiencies of 30% alone. Predicted epidemic trajectories for the unmitigated scenario (A), range of detection rates (B), and range of NPI efficiencies (C). Results from 100 simulations are shown in A with the black line representing the median number of cases. Shaded regions represent the 95% confidence intervals around the median in panels B and C. All simulations began on the date of the first positive imported case in Madagascar, March 20, 2020. The y-axis is plotted on the log10-scale.

First, case definitions in Madagascar may be stricter than elsewhere. In June 2020, about 12% of suspected cases tested were confirmed by RT-PCR in Madagascar compared to a positivity rate of less than 5% in Europe and the US over the same period [30,31]. This suggests strict criteria for test eligibility (e.g. requiring symptoms and known positive contacts) is being used in Madagascar and many cases could potentially be missed. Further, strict diagnostic criteria may be a result of the original testing policy of Madagascar early in the epidemic, which required that suspected cases first test positive with antibody-based rapid detection tests (RDTs) before being confirmed through RT-PCR tests. The low probability of detection of antibodies early in the infectious period (i.e., the first week of symptoms) and the high rate of false-negatives with RT-PCR later in the infectious period (i.e., after 7-10 days post symptom onset) [32] create a short window for detection.

Second, the Madagascar health system itself is only receiving a portion of symptomatic COVID-19 cases. The proportion of asymptomatic COVID-19 cases is estimated to be between 40–45% [33]. In Madagascar, surveys indicate that 40.2% of people with respiratory infection symptoms seek healthcare [34], implying that over 50% of symptomatic COVID-19 infections may not even enter a public health facility. For COVID-like symptoms, this rate could be much lower given the stigma associated with the disease [11]. The combination of the high proportion of asymptomatic cases and low health-seeking behaviors suggest that, even if health centers test 50% of symptomatic COVID-19 cases attending a health facility, this would detect less than 25% of symptomatic cases.

Finally, there is limited diagnostic testing capacity in Madagascar, with RT-PCR testing available in 5 laboratories across 3 major cities. It is unlikely that health facilities in rural areas of the country, where nearly 50% of the population lives, are testing such a high percentage of cases. We can account for all of these factors to estimate an upper bound of detection rates for Madagascar (strict case definitions (0.3) × low proportion symptomatic (0.45) × low healthcare-seeking behaviors (0.2) × limited testing infrastructure (0.25)) to reasonably explain a detection rate in Madagascar of 1% or lower.

## Reduced Transmission

A reduction in transmission rates of 30%, relative to an unmitigated scenario, can also explain the daily case report rates of COVID-19 in Madagascar (Fig. 1C). This reduction could be the result of NPI policies put in place in Madagascar or of innate characteristics affecting the epidemiology of COVID-19 (e.g. baseline contact patterns, climate, etc.). NPIs were implemented within three days after the first confirmed imported case of COVID-19 in the country (Table 1), the majority of which focused on restricting intercity travel on roadways and included lockdowns in population centers. In contrast, the UK instituted a partial-lockdown on March 23, 2020, 52 days after the first confirmed case in the UK on January 31, 2020. The road system of Madagascar is highly fragmented, with most travel on a limited number of paved national roads that run North-South through the capital. Restricting travel on these roads has the potential to be highly effective in reducing human mobility in Madagascar, and therefore the spread of COVID-19. Further, most travel involves major population centers, particularly the capital city, Antananarivo [35]. These cities had much more stringent NPIs put in place early in the epidemic, including city-wide lockdowns and curfews (Table 1), and the targeted lockdown of these population centers could have reduced spread to the rest of the country. While mobility data is not available for Madagascar, other SSA countries have reported reductions in mobility ranging from 1.4% in Zambia to 19% in Senegal compared to pre-NPI levels [36]. With a sparse road network that is well regulated in Madagascar, 30% represents an obtainable reduction in contact rates.

Because NPIs were implemented early in the epidemic, their effects on transmission cannot be disentangled from baseline contact patterns in the country, which may be lower than those of Europe or the US. Nearly half (47.73%) of the Malagasy population lives in rural areas, and most of the country is over 3 hours from a population center with more than 50,000 people [37]. Therefore, baseline contact patterns in the rural areas of Madagascar may be reducing disease spread in a way that is unidentifiable from the effects of NPIs.

## Which path is Madagascar on?

The evidence presented here provides no indication that the epidemiology (e.g. *R*_0_) of COVID-19 is fundamentally different in a fairly typical SSA country than elsewhere. We demonstrate that the current trend in reported cases in Madagascar can be explained by its early stage in the epidemic, combined with low detection rates and lower contact rates from NPIs (Fig. 2A). Understanding how much of the discrepancy between predicted and reported case burdens is due to low detection rates or NPIs has enormous implications for our expectations regarding the ‘true’ burden of COVID-19 in Madagascar. For this, we explored different combinations of detection rates and NPI efficacy that explain the observed trend in reported cases, together with associated predictions of epidemic morbidity and mortality burdens (Figure 2). If the low number of reported cases is due primarily to a low detection rate, we predict over 13 million people will be infected with the virus if NPIs are not in place (Fig. 2C,D), imposing a huge burden on an already weakened health system. On the other hand, if the low number of cases is due to a reduction in contact patterns, the model predicts a lower total burden of approximately 8 million people infected with the virus (Fig. 2C,D). If NPIs are driving these contact patterns and are responsible for the lower-than-expected case burden, many cases and deaths are being averted, but the lifting of these restrictions is very likely to lead to an uncontrolled outbreak.

**Figure 2.**
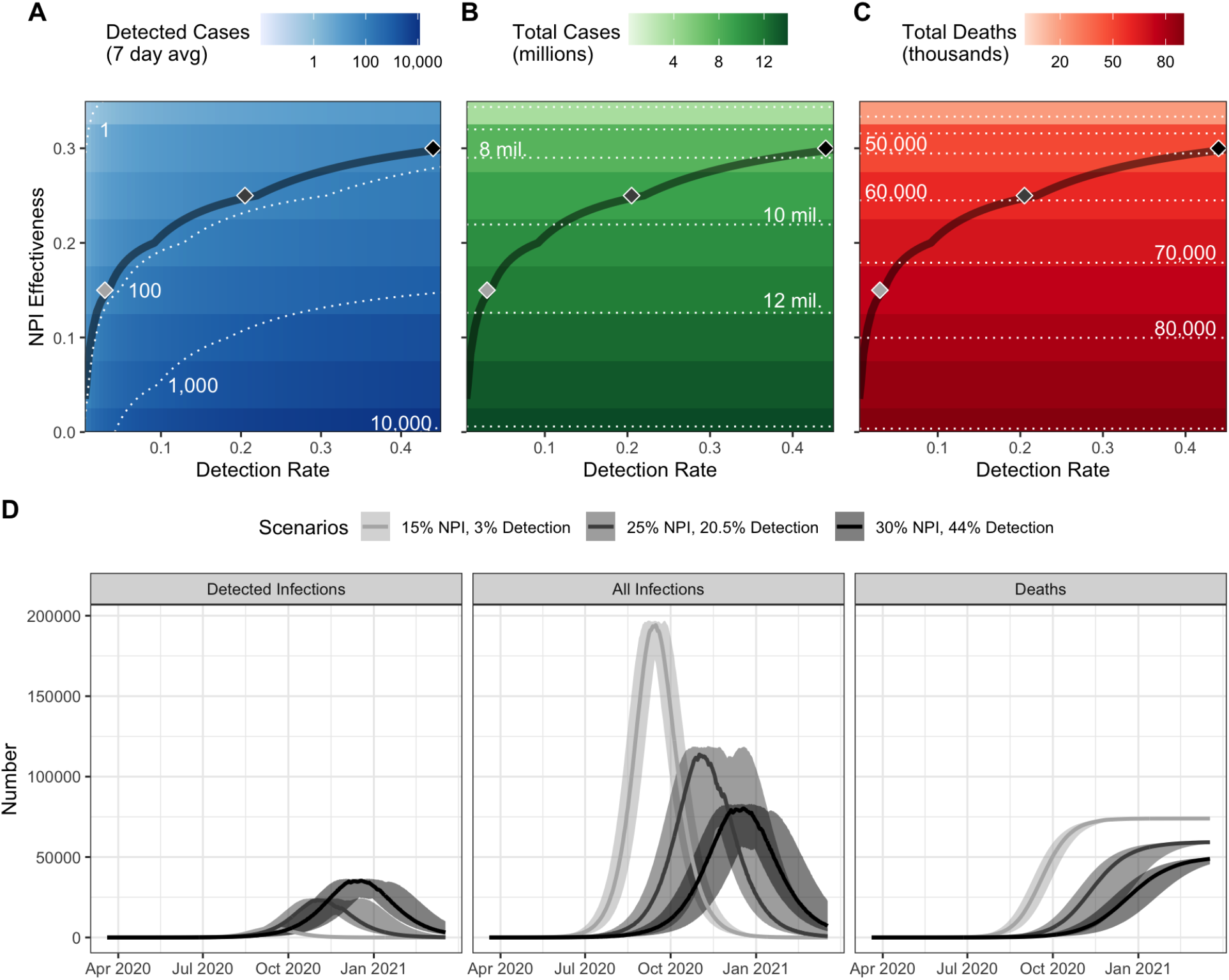
Multiple combination of factors explain the low number of reported COVID-19 cases in Madagascar, with important implications for predictions of epidemic dynamics. Predicted A) daily reported cases (7 day average) as of June 22 2020, B) total infection burden after one year, and c) total fatality burden after one year in Madagascar at different combinations of NPI effectiveness and detection rates. The contour line corresponds to a parameter space where the median number of predicted cases from 25 simulations equals the daily reported cases (7 day average) on June 22 (71.71 cases). Shaded diamonds correspond to specific scenarios explored in panel D, illustrating the dynamics of detected infections, all infections, and cumulative deaths over the first year of the epidemic.

Although the global epidemic began several months ago, current infectious disease models for Madagascar and other SSA countries rely on limited data, resulting in disparate predictions. Pearson et al. (2020) predicted a similar epidemic size for Madagascar as our model in an unmitigated scenario, with 75% of the population and nearly 100,000 deaths. In contrast, an analysis led by the WHO [12] predicted a total case burden nearly one third of this size (26% of the population) and only a fraction of COVID-19 related deaths (1,500). This study assumed that the regional particularities of SSA will decrease disease transmission and fatality rates based on country-specific proxies for these factors, such as climate, transportation networks, and contact matrices. Importantly, this study only considered *reductions* in transmission via reduced risks of exposure, with a maximum of 2.6% of the population of Madagascar at risk of exposure at any one time. While socio-ecological context is necessary to understand disease transmission, our exercise suggests that the difference between reported and predicted case burdens in SSA can be just as easily explained by accounting for low detection rates and NPIs that reduce interpersonal contact.

## Conclusion

We do not currently have enough evidence to suggest that the epidemiology of COVID-19 is different in Madagascar than elsewhere. The low number of reported cases can be explained by low detection rates, late introduction, and early and effective implementation of NPIs. In contrast to the theory of a salutary epidemiology, each of these explanations is supported by strong anecdotal evidence (Table 2). As lockdowns are gradually lifted, other NPIs, such as handwashing and social distancing, should be implemented to avoid a rapid growth in cases. The public health system should remain prepared for an outbreak, with a peak of infections expected between August and December depending on the transmission scenario (Fig. 2D). The COVID-19 epidemic could become the leading public health problem in Madagascar in 2020, causing nearly twice as many deaths as are attributed to diarrheal disease [38]. It is important, therefore, to conduct clinical trials and continually improve access to health care. If NPIs remain in place at levels seen during the first months of the epidemic, the model suggests that this could prevent over 50,000 COVID-19 related deaths in Madagascar.

**Table 2.** Summary of evidence supporting or opposing three possible explanations for the low number of reported cases of COVID-19 in Madagascar.

|  | Supporting | Opposing |
| --- | --- | --- |
| **Low detection rates** | - High proportion of asymptomatic cases[33]<br>- Strict testing criteria<br>- Low healthcare seeking rates for acute respiratory infections[34]<br>- Diagnostic practices that limit the window of detection | - Recent evaluation of health system preparedness via the International Health Regulations meant health systems were on high alert for an outbreak [20] |
| **Epidemiological differences** | - Trained-immunity due to vaccinations or high prevalence of endemic disease could increase population's resistance to infection [14]<br>- Transmission rate may be lower in sparsely populated areas [13]<br>- Virus survival is lower in humid, warm environments | - Limited role for climate during pandemic phase of the outbreak [16]<br>- Past influenza outbreaks were not limited by sparse transport networks in SSA [17] |
| **Early and effective NPIs** | - Lockdown in population centers implemented three days after first imported case<br>- Limiting travel on fragmented paved road network can easily disrupt within-country movement |  |

## Data Availability

All data is from the open dashboard on the Africa Centers for Disease Control and Prevention website.

## Funding and Acknowledgements

MVE was supported by a Graduate Research Fellowship (National Science Foundation). We thank Tanjona Ramiadantsoa and Laura Cordier for their insights and comments on earlier drafts of this manuscript.

